# Landing rates of *Aedes aegypti* and *Culex quinquefasciatus* in Northern Ecuador: Human Landing Catch Study, 2021-2022

**DOI:** 10.1101/2025.03.12.25323448

**Authors:** Patricio Ponce, Ian Pshea-Smith, Varsovia Cevallos, Oscar Suing, Andrés Carrazco-Montalvo, Amy C. Morrison, Josefina Coloma, Joseph N. S. Eisenberg

## Abstract

The landing patterns of mosquitoes are critical for disease prevention given their roles as vectors of numerous pathogens, especially given documented variability in their patterns across different geographies and contexts. In Ecuador, two of the most important vectors associated with human arboviruses infection, *Aedes aegypti* and *Culex quinquefasciatus*, are abundant. Ascertaining the patterns of these mosquitoes’ biting behaviors (and through this, their potential to transmit pathogens) is essential to better tailor public health interventions and thereby prevent the spread of arboviral diseases. From June 2021 to July 2022, we performed human landing collections of mosquitoes in two sites within Northern Ecuador, capturing mosquitoes in protected and unprotected settings, across the day’s span. We captured 18,280 mosquitoes, 95.51% were *Cx. quinquefasciatus* and 1.8% were *Ae. aegypti*. The results of this study reinforce traditional diel landing patterns and crepuscular patterns for *Aedes aegypti* and *Culex quinquefasciatus*, respectively. We emphasize the importance of elucidating landing dynamics in varied and unique contexts, as established patterns and new dynamics are not necessarily consistent across geographies. Finally, we describe differences in protected versus unprotected collection events, with vector species being more likely to land on protected collectors, of importance for future landing catch studies in disease-endemic areas.

## Introduction

Mosquitoes are important disseminators of disease worldwide, spreading pathogens such as dengue virus, West Nile virus, chikungunya virus and Zika virus [1–3]. These viruses and key mosquito vectors are endemic throughout Latin America, including Ecuador, which reported more than 16,000 and 23,000 dengue fever cases in 2022 and 2023, respectively [4–6].

In Ecuador, two of the most important vectors associated with human arboviruses infection, *Aedes aegypti* and *Culex quinquefasciatus* are abundant. Ascertaining the patterns of these mosquitoes’ biting behaviors (and through this, their potential to transmit pathogens) is essential to better tailor public health interventions and thereby prevent the spread of arboviral diseases [7,8].

These biting behaviors vary within and across genera and species, and are inherently complex, necessitating evaluation and re-evaluation of key vector’s behaviors in new and varied contexts, especially in this time of global climate change [9–12]. Of particular note, diel landing patterns, specifically of *Ae. aegypti*, have been described to be shifting in recent years – therefore, identifying and verifying when this and other arbovirus vectors land throughout the day in distinct contexts is of growing importance [13–15].

We collected mosquitoes using the human landing catch (HLC) method in two locations across a year’s span, because other traps fail to accurately and holistically assess which mosquitoes seek and feed upon human hosts [16–18]. Moreover, to understand exposure, HLC methods facilitate monitoring hourly abundance patterns and most accurately approximate the per-person-time biting rates of mosquitoes, in addition to capturing diel patterns that other methods may miss [16,17].

Our study describes diurnal patterns of *Ae. aegypti* and *Cx. quinquefasciatus* host seeking behaviors in northwestern Ecuador and compares two human bait methodologies that use protected and unprotected hosts.

## Methodology

### Study Setting

Mosquito collections were carried out in in two communities in northwestern Ecuador, within Esmeraldas Province. One location serves as a regional hub for commerce and travel (Borbón). This community has between 8,000 and 9,000 residents, and was previously described [19]. The second location is a riverine village with less infrastructure (Santa María). Santa María has a population between 500 and 600 individuals as described by Lee et al. in 2021 [20]. A single dwelling, where owners and inhabitants provided permission for the collections, was selected in each community to carry out both unprotected and protected human landing collections (HLC). Unprotected collections were carried out by individual collectors with exposed lower limbs while sitting immediately next to the dwelling and alongside the road. In contrast, protected collections refer to a double net method where collections were carried out within a semi-enclosed space and covered using a canopy of fine netting. This canopy was approximately 2.5 high and 2.5 meters in diameter and was enclosed from the top extending down to 0.5 meters in height. When studying *Aedes aegypti*, performing indoor captures are important given its indoor resting and biting habits shown across other contexts [18,21,22]. In this study, we were limited to outdoor collections – the unprotected approach as an alternative sought to examine both if partially protecting the collectors resulted in differing landing rates, and if it could serve as a potential proxy to study indoor landing dynamics when an indoor capture isn’t possible.

### Mosquito Collection

Monthly mosquito collections were conducted between June 2021 and July 2022 during the second week of each month. Each monthly collection consisted of three daily collection periods repeated daily over four consecutive days. The three collection periods were 05:00 to 08:00 (morning), 1100 to 1400 (midday) and 1700 to 2000 (evening). June 2022 was skipped due to civil unrest. A pilot week with nocturnal collections (after 2000) was performed, however due to no *Aedes aegypti* being collected and prior knowledge highlighting diurnal landing of this species, this study employed only diurnal collections.

At least six individuals participated in mosquito collection, alternating between protected and unprotected methods every hour and rotating between locations (Borbón and Santa María) each month to minimize collector bias. Collections were conducted using handheld, Prokopak® aspirators. Mosquitoes were aspirated upon landing on the exposed lower legs of the collectors and transferred to collection vials until the collection period ended. After the collection period was completed, mosquitoes were placed at −80 for at least one hour before identification. Identification was performed to the genus level by entomology technicians before trained entomologists speciated the intact specimens.

### Covariate Data

Collectors used electronic data loggers to assess temperature (□), humidity (%) and wind speed (meters/s). Each were recorded every fifteen minutes during collection periods, in both Borbón and Santa María. We calculated the mean value for each variable during each hourly period. To estimate daily rainfall for the collection period we used NASA’s Integrated Multi-Satellite Retrievals for global precipitation measurements for each site [23]. For analyses, we calculated running sums and means of precipitation from the previous week and previous two weeks.

### Statistical Analyses

Data analysis and visualizations were performed using RStudio 2023.09.1 Build 494, and Microsoft Excel Version 2309 (Build 16827.20278). All code and data are available via GitHub (https://github.com/IanPsheaSmith/Ecuador_LandingCatch). Missing observations for temperature, humidity and wind speed were imputed; imputation was based on mean values of the missing observations nearest temporal neighbors in its location. For example, a missing temperature at 0700 in Borbón would be imputed based on the mean temperature of 0600 and 0800 that same day in Borbón. Missingness for an entire day would be imputed based on the mean values for each corresponding time from the previous and subsequent days during that month.

Total counts of the identified mosquitoes were stratified by location and protected/unprotected collector status (Table 1). Means and medians for temperature, humidity, wind speed and daily precipitation when female *Ae. aegypti* were absent or present were determined for each hourly period and then compared using two-sample Wilcoxon rank-sums tests (S1 Table). This was then repeated for the absence and presence of female *Cx. quinquefasciatus*.

**Table 1.** Counts of all captured mosquitoes by location, collector protected/unprotected status and sex. Mosquitoes identified to the genus but not to the exact species are shown as *sp*. *One female *Culex* mosquito had intact location information but was missing protected/unprotected capture status.

|  | Location |  |  |  | Capture Setting |  |  |  |
| --- | --- | --- | --- | --- | --- | --- | --- | --- |
|  | Borbón |  | Santa María |  | Protected |  | Unprotected |  |
|  | Females | Males | Females | Males | Females | Males | Females | Males |
| ***Aedes*** |  |  |  |  |  |  |  |  |
| *aegypti* | 121 | 164 | 15 | 20 | 105 | 159 | 31 | 25 |
| *angustivittatus* | 4 | 0 | 1 | 0 | 1 | 0 | 4 | 0 |
| *pertinax* | 3 | 0 | 0 | 0 | 1 | 0 | 2 | 0 |
| *scapularis* | 14 | 0 | 0 | 0 | 5 | 0 | 9 | 0 |
| *serratus* | 22 | 3 | 0 | 0 | 7 | 0 | 15 | 3 |
| *taeniorhynchus* | 1 | 0 | 0 | 0 | 1 | 0 | 0 | 0 |
| ***Anopheles*** |  |  |  |  |  |  |  |  |
| *albimanus* | 3 | 0 | 0 | 0 | 0 | 0 | 3 | 0 |
| *calderoni* | 22 | 0 | 1 | 0 | 21 | 0 | 2 | 0 |
| *apicimacula* | 0 | 0 | 1 | 0 | 1 | 0 | 0 | 0 |
| ***Coquillettidia*** |  |  |  |  |  |  |  |  |
| *nigricans* | 0 | 1 | 0 | 0 | 0 | 1 | 0 | 0 |
| *lynchi* | 2 | 0 | 0 | 0 | 1 | 0 | 1 | 0 |
| *nigricans* | 117 | 47 | 1 | 0 | 83 | 45 | 35 | 2 |
| sp. | 10 | 2 | 0 | 0 | 6 | 2 | 4 | 0 |
| ***Culex*** |  |  |  |  |  |  |  |  |
| *nigripalpus* | 40 | 10 | 51 | 24 | 64 | 32 | 27 | 2 |
| *quinquefasciatus* | 310 | 404 | 5254 | 10568 | 4605 | 8928 | 959 | 2044 |
| *spissipes* | 7 | 0 | 4 | 0 | 7 | 0 | 4 | 0 |
| *vomerifer* | 7 | 0 | 0 | 0 | 6 | 0 | 1 | 0 |
| sp. | 31 | 6 | 541 | 203 | 543* | 203 | 28* | 6 |
| ***Johnbelkinia*** |  |  |  |  |  |  |  |  |
| *ulopus* | 0 | 0 | 1 | 0 | 0 | 0 | 1 | 0 |
| sp. | 0 | 0 | 2 | 0 | 2 | 0 | 0 | 0 |
| ***Limatus*** |  |  |  |  |  |  |  |  |
| *durhamii* | 2 | 1 | 0 | 0 | 2 | 0 | 1 | 0 |
| ***Lutzia*** |  |  |  |  |  |  |  |  |
| *allostigma* | 0 | 0 | 1 | 0 | 1 | 0 | 0 | 0 |
| ***Mansonia*** |  |  |  |  |  |  |  |  |
| *titillans* | 16 | 0 | 0 | 0 | 12 | 0 | 4 | 0 |
| *amazonensis* | 93 | 1 | 0 | 0 | 58 | 0 | 35 | 1 |
| *indubitans* | 16 | 0 | 0 | 0 | 10 | 0 | 6 | 0 |
| *pseudotitillans* | 7 | 0 | 0 | 0 | 5 | 0 | 2 | 0 |
| *wilsoni* | 68 | 0 | 0 | 0 | 30 | 0 | 38 | 0 |
| ***Psorophora*** |  |  |  |  |  |  |  |  |
| *circumflava* | 3 | 0 | 1 | 0 | 3 | 0 | 1 | 0 |
| *ferox* | 4 | 0 | 1 | 0 | 4 | 0 | 1 | 0 |
| *albipes* | 7 | 0 | 1 | 0 | 6 | 0 | 2 | 0 |
| *champerico* | 12 | 0 | 1 | 0 | 9 | 0 | 4 | 0 |
| sp. | 1 | 0 | 0 | 0 | 1 | 0 | 0 | 0 |
| ***Trichoprosopon*** |  |  |  |  |  |  |  |  |
| *hyperleucus* | 0 | 0 | 1 | 0 | 1 | 0 | 0 | 0 |
| sp. | 1 | 0 | 1 | 0 | 0 | 0 | 2 | 0 |
| ***Wyeomyia*** |  |  |  |  |  |  |  |  |
| sp. | 0 | 0 | 3 | 0 | 3 | 0 | 0 | 0 |
| **Total** | **944** | **639** | **5882** | **10815** | **5604** | **9370** | **1222** | **2083** |

### Logistic Regression

To discern if hourly landing rates varied, we constructed logistic regression models based on the presence/absence of each species [24]. Models were chosen using forward stepwise regression with dredging, based on Akaike’s Information Criteria – this was done to form the most parsimonious model. For the forward stepwise regression, we employed the R package MuMIn [25].

In addition to collection time, all covariates (temperature, humidity, wind speed and rainfall [daily, weekly lag and two weeks lag]) were included in the stepwise regression process, with three categorical dummy variables: protected/unprotected collector status, location (Borbón/Santa María), and collection month. The final logistic regression models are displayed in Table 2, showing odds ratios only for statistically significant (α = 0.05) variables that were included in the final model based on stepwise regression.

**Table 2.** Logistic regression model examining the presence and absence of female *Aedes aegypti* and female *Culex quinquefasciatus* per hourly period. The probability modeled is female *Ae. aegypti* ≥ 1 or female *Cx. quinquefasciatus* ≥ 1 in any given hourly period. Note that hourly odds shown here may not coincide with visual trends in Figure 2, as these models adjust for other covariates and model only presence, so do not account for occasional events where 2 or more of the species were present. Sample interpretation: The odds of at least one female *Culex quinquefasciatus* mosquito being captured in one hourly period were 16.13 times higher in Santa María than in Borbón, holding all other included variables constant. NS: Variable is not significant at an alpha level of 5%. ∼ Variable was excluded from the final model – month and humidity were excluded based on stepwise regression for *Ae. aegypti*, and temperature was excluded based on stepwise regression for *Cx. quinquefasciatus*. Rainfall (including various lags) was excluded from both models based on stepwise regression. * The month of June was skipped in 2022 due to civil unrest.

| Covariate | Odds Ratio (95% confidence interval) |  |
| --- | --- | --- |
|  | *Aedes aegypti* | *Culex quinquefasciatus* |
| Collection Time |  |  |
| 0500-0600 | Reference | Reference |
| 0600-0700 | NS | NS |
| 0700-0800 | NS | 0.19 (0.09, 0.40) |
| 1100-1200 | 4.21 (1.10, 16.16) | 0.24 (0.10, 0.62) |
| 1200-1300 | 4.45 (1.13, 17.55) | 0.23 (0.08, 0.67) |
| 1300-1400 | NS | 0.25 (0.09, 0.74) |
| 1700-1800 | 5.14 (1.41, 18.70) | NS |
| 1800-1900 | NS | 2.60 (1.42, 4.78) |
| 1900-2000 | 0.15 (0.02, 0.97) | NS |
| Protected/Unprotected |  |  |
| Protected | Reference | Reference |
| Unprotected | 0.57 (0.35, 0.94) | 0.23 (0.16, 0.33) |
| Location |  |  |
| Borbón | Reference | Reference |
| Santa María | 0.12 (0.06, 0.23) | 16.13 (10.35, 25.13) |
| Month |  |  |
| June 2021 | ~ | Reference |
| July | ~ | NS |
| August | ~ | 2.90 (1.37, 6.16) |
| September | ~ | 3.55 (1.68, 7.50) |
| October | ~ | 5.57 (2.77, 11.18) |
| November | ~ | NS |
| December | ~ | NS |
| January 2022 | ~ | 3.41 (1.58, 7.37) |
| February | ~ | 8.71 (4.50, 16.86) |
| March | ~ | 3.38 (1.64, 6.99) |
| April | ~ | 3.88 (1.82, 8.30) |
| May | ~ | NS |
| June | * | * |
| July | ~ | NS |
| Temperature (°C) | 1.17 (1.05, 1.31) | ~ |
| Humidity (%) | ~ | 1.06 (1.02, 1.10) |

### Ethics statement

The protocol for this study was reviewed and approved as part of an arboviral surveillance study. This work was approved by the Universidad San Francisco de Quito (2017-159M) and the University of Michigan (HUM00140967) ethics review boards, and was also reviewed and approved by the Ecuadorian Ministry of Health (Ministerio de Salud Publica) at the local and national levels (MSPCURI000237-2). All adult participants provided written informed consent.

## Results

### Total Mosquitoes

A total of 18,280 mosquitoes were collected between June 2021 and July 2022. Of these, 6,826 were female, and 11,454 were males. The most abundant species in both sites was *Cx. quinquefasciatus* (17,460 of 18,280, 95.51%). *Aedes aegypti* comprised only 1.8% of the mosquitoes captured. Mosquitoes were more abundant in Santa María (n=16,6895, 91.33%), than Borbón (1,382, 7.56%); however, most of the *Ae. aegypti*, 88.97% of females (*X^2^* = 162, *df* = 1, *P* < 0.001) and 89.13% of males were collected in Borbón (*X^2^* = 222, *df* = 1, *P* < 0.001). Additionally, more mosquitoes were collected using the protected approach than the unprotected approach, with 14,975 (81.94%) being collected in a protected setting compared to only 3,301 in an unprotected setting. Counts of both target species were significantly higher using protected versus unprotected collections - 264 *Ae. aegypti* were captured in the protected setting compared to 54 in the unprotected setting (*X^2^* = 274.72, *df* = 1, *P* < 0.001), and 13,533 *Cx. Quinquefasciatus* collected in the protected setting compared to 3,003 in the unprotected setting (*X^2^* = 13408, *df* = 1, *P* < 0.001).

### Temporal Patterns of Female *Aedes aegypti* & *Culex quinquefasciatus*

One female *Ae. aegypti* and seventeen female *Cx. quinquefasciatus* could not be matched to their collection times, and were excluded from temporal analyses and modelling. Diurnal landing activity of *Ae. aegypti* females was highest during three midday hours (61 or 45.19% of 135) and evening (48, 35.55%) than in the morning (26, 19.26%) (Fig 2). This difference was found to be statistically significant using a chi-squared test, with a subsequent p-value of 0.001 (*X*-squared = 13.911, df = 2). In contrast, *Cx. quinquefasciatus* females had higher landing rates in the evening hours (4,084 or 73.63% of 5,547) and morning hours (1,336, 24.08%) with only 127 (2.28%) being captured during midday (*X^2^* = 7531.5, *df*=8, *P* < 0.0001).

Female *Ae. aegypti* abundance was highest during the June 2021, January 2022 and April 2022 collections, with 20, 19 and 22 female mosquitoes being captured in these respective months (Fig 1). No clear monthly pattern was elucidated for *Ae. aegypti* (Fig 1). For female *Cx. quinquefasciatus,* abundance was highest in February of 2022, with 61 (1.1% of all female *Cx. quinquefasciatus*) captured in Borbón and 1,218 (23.18%) captured in Santa María, for a total of 1,251 (22.48%) in February (Fig 1).

**Figure 1.**
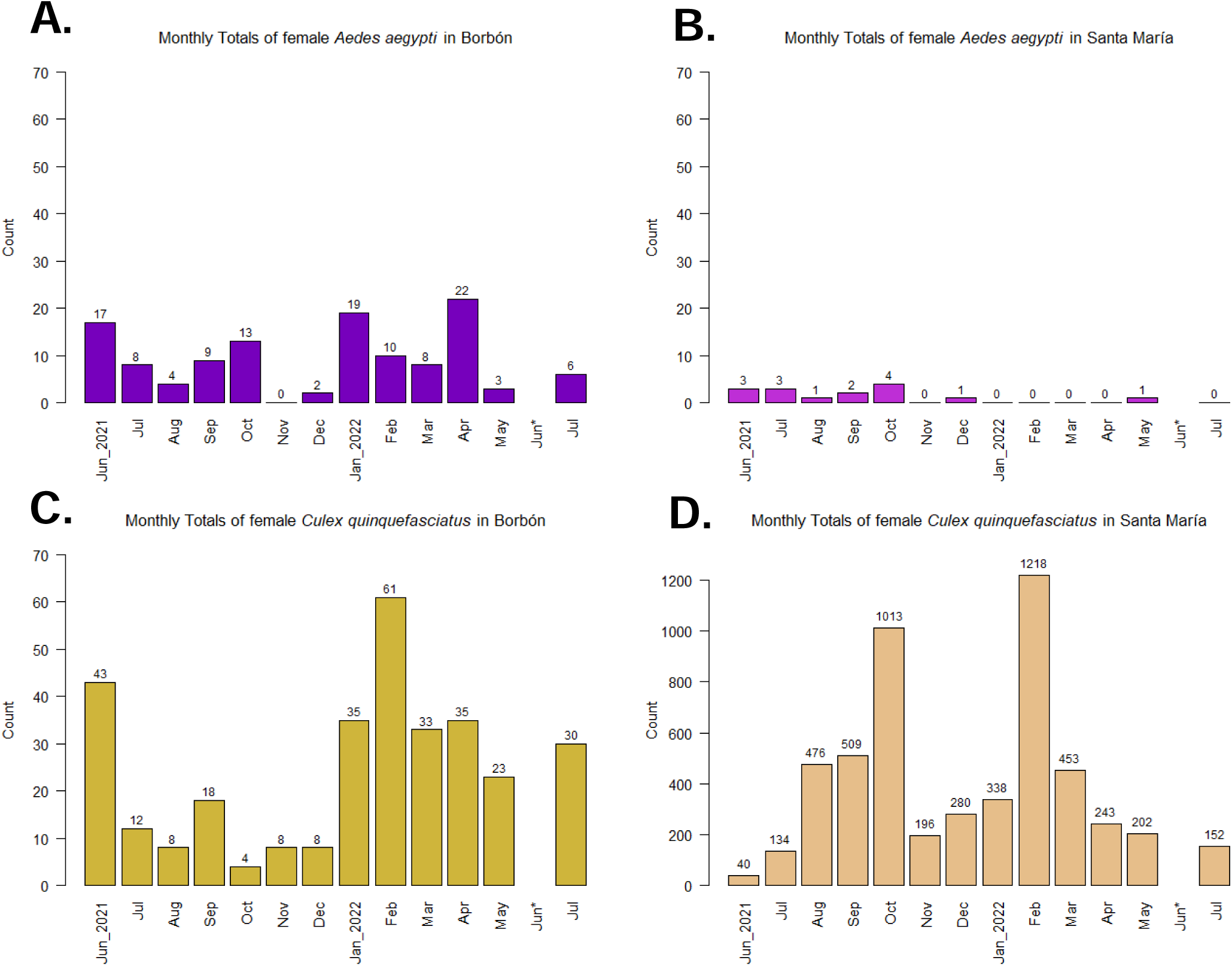
Monthly counts of female *Aedes aegypti* and *Culex quinquefasciatus* in Borbón (A & B) and Santa María (C & D). Six *Ae. aegypti* females and 182 *Cx. quinquefasciatus* females were missing monthly labels and are excluded from this figure. *Note that June 2022 was skipped due to civil unrest and note that the scale of the y-axis for D (*Cx. quinquefasciatus* in Santa María) differs.

### Covariate Associations

Temperature, humidity and wind speed for when females of either species were present or absent were statistically different ( = 0.05; Wilcoxon Rank-Sum tests), while rainfall did not differ significantly (S1 Table). Humidity and wind speed were excluded in the final iteration of the logistic regression models due to a combination of a lack of correlation in the model or a worsening of model fit criteria, perhaps indicating other variables such as location or time are driving these associations.

The odds of capturing at least one female *Aedes aegypti* in an hourly period during 1100-1200, 1200-1300, and 1700-1800 were statistically higher when compared to the reference period of 0500-0600 while holding all other variables in the model constant (Table 2). The odds of capturing at least one *Ae. aegypti* female in an hourly period for an unprotected collector were 0.57 (95% CI: 0.35, 0.94) times lower than for collecting mosquitoes while protected, holding the other modelled variables constant, and were 0.12 (0.06, 0.23) times lower at the site in Santa María than in Borbón. Temperature was positively associated with the presence of at least one female *Ae. aegypti* in an hourly period; for each one-degree Celsius increase in temperature, the odds of finding at least one increased by 17% (95% CI: 5%, 31%).

The odds of capturing at least one female *Cx. quinquefasciatus* mosquito during the hourly periods of 0700-0800, 1100-1200, 1200-1300 and 1300-1400 were lower in comparison to the reference time of 0500-0600, while the odds were higher during 1800-1900 with an odds ratio of 2.60 (1.42, 4.78) (Table 2). The odds of finding at least one female *Cx. quinquefasciatus* mosquito for an unprotected collector were 0.23 (0.16, 0.33) times lower than finding them for a protected collector. Comparing the odds of capture at the two sites, the odds of finding at least one in the site at Santa María were 16.13 (10.35, 25.13) times higher than in the site at Borbón. Compared to June 2021, the odds of capturing at least one female *Cx. quinquefasciatus* mosquito during an hourly period were higher during August, September, October, January, February, March and April, and were not significantly different during the other months. Humidity was also positively associated with both the presence and the counts of *Cx. quinquefasciatus*; for each one-unit increase in humidity, the odds of finding at least one is multiplied by 1.06 (1.02, 1.10).

## Discussion

Both female *Aedes aegypti* and female *Culex quinquefasciatus* exhibited clear diel activity patterns that are consistent with patterns reported previously in the literature [13,14,26–29]. *Ae. aegypti* activity was highest during mid-day and showed no tendencies toward nocturnal activity despite artificial light being available in both sites. *Cx. quinquefasciatus* showed clear crepuscular activity. Also notable was that our protected collections were more effective than unprotected collections, which may reflect that the structure surrounding the netted collector simulated an indoor environment, where both species have available resting sites. This finding contributes novel insight regarding the efficacy of protected collections in a unique geographic context.

Historically, *Ae. aegypti* mosquitoes tend to maintain a diurnal landing period, and herein were also determined to bite across the day with a mid-day preference [13,14,26,30]. However, recent work and discussions within the field have highlighted concerns that these mosquitoes may be shifting into nocturnal biting habits, especially under dynamics of increased artificial lights at night [13,31]. For example, Chadee described nocturnal biting dynamics and a correlation with light intensities and mosquito landing patterns, with implications of increasing nocturnal activity in an urbanizing world [31]. Our findings for *Ae. aegypti,* while they do not incorporate luminosity or measure landing past 20:00, still show consistent diel landing rates, and the odds of capturing a mosquito past 19:00 were significantly lower than the reference time; this is of importance for vector-borne disease control and prevention, as this diel patternicity presents a clear barrier. Preventative methodologies such as bed nets or heading indoors at night cannot prevent predation by *Ae. aegypti* within this context.

In contrast, *Cx. quinquefasciatus* exhibited distinct crepuscular landing patterns; their odds of landing were lower in the midday when compared to the morning, and were much higher for the evening time periods (Fig 2, Table 2). These patterns are consistent with established crepuscular and nocturnal behaviors for this vector in other contexts [27–29,32,33]. Considering this pattern exhibited by *Cx. quinquefasciatus* in this context, akin to *Ae. aegypti*, disease transmission prevention may necessitate approaches such as pesticide application or population control mechanisms such as larval source elimination; while this species did have an evening preference, human inhabitants are still active in the later evening. Additional studies into the nocturnal patterns of this mosquito in this context may be beneficial to compare with these findings.

**Figure 2.**
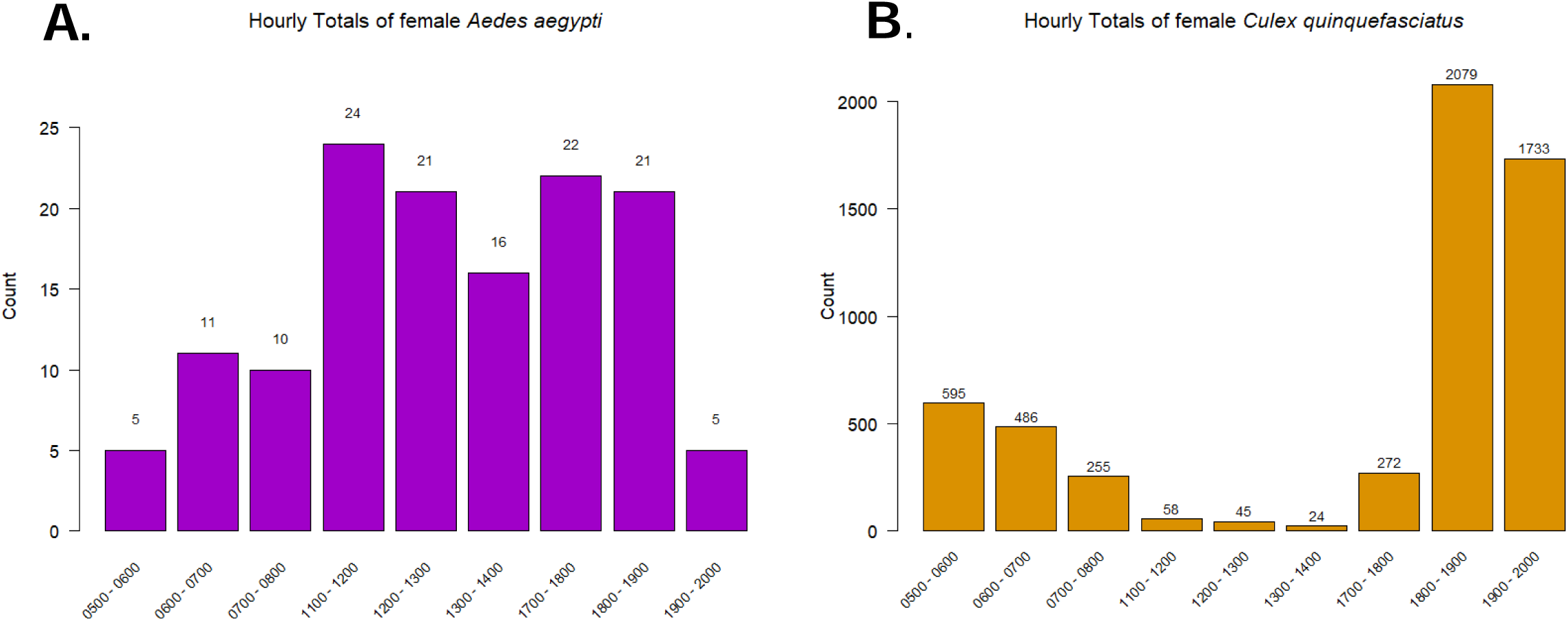
Counts of female *Aedes aegypti* (A) and female *Culex quinquefasciatus* (B) by hourly period. One female *Ae.* aegypti and seventeen *Culex* quinquefasciatus females were captured outside of these periods and are excluded from this figure. Note that the scale of the y-axis differs.

We also herein elucidate statistically significant differences in the odds of both species being present for protected versus unprotected collection events. For *Ae. aegypti*, the odds of capturing at least female mosquito were 0.57 (95% CI: 0.35, 0.94) times lower for unprotected collectors in comparison to protected collectors, and were even lower for *Cx. quinquefasciatus* (OR: 0.23, 95% CI: 0.16, 0.33). Importantly, this pattern persists while adjusting for other covariates, such as temperature and seasonality. This finding suggests that partial protection was not beneficial in preventing biting from either mosquito species in this context. Domestic versus peridomestic (potentially, partially protected versus unprotected) landing rates vary by species and context, with multiple studies demonstrating that varied preferences emerge across genera and species [14,34,35]. For instance, a review of *Ae. aegypti* biting rates identified six studies with higher biting activity outdoors, and only one study with a higher indoor rate [18,31,36–38]. Of note however, robust work has shown large abundances of this species resting indoors, across broad geographies such as Mexico, Trinidad and Thailand [39–41]. Studies on *Culex quinquefasciatus* presented similarly divergent findings, with studies in Nigeria and Benin showing no indoor/outdoor preference while multiple studies in Indonesia displayed a higher indoor preference, while this species is also recognized as being abundant in indoor resting studies [42–46]. Even small degrees of indoor landing are of importance, as this behavior implies routine exposure to the species within the home, where families may spend significant amounts of time. We emphasize the importance of regionally and locally specific studies to identify specific landing trends in unique and novel geographies, to better elucidate these patterns.

We also describe several covariate associations with the odds of each species being present. These dynamics have been reviewed intensively elsewhere, and our results of temperature and humidity being of importance align with our understanding of mosquito’s ectothermic biology [47,48]. Landing behaviors are inherently complex, with multiple intersecting factors that impact and alter them. While temperature, humidity, light, weather patterns and other environmental or climactic factors play an innate role, in this particular context, the location, protected/unprotected status, hourly and monthly variables were the most impactful from a modeling perspective [13,47,49,50]. However, our findings should be contextualized and further work can be performed to better elucidate the patterns we found. For example, unmeasured variables such as the land use surrounding each site or factors such as human activity and the density of the population around these specific points may alter mosquito behaviors [51–53].

The results of this study reinforce traditional diel landing patterns and crepuscular patterns for *Aedes aegypti* and *Culex quinquefasciatus*, respectively. We wish to highlight the importance of elucidating landing dynamics in varied and unique contexts, as established patterns and new dynamics are not necessarily consistent across geographies. Finally, we describe differences in protected versus unprotected collection events, with vector species being more likely to land on protected collectors, of importance for future landing catch studies in disease-endemic areas. This study provides novel evidence that can be used to adapt vector control strategies to specific local contexts.

## Data Availability

All code and data are available via GitHub

https://github.com/IanPsheaSmith/Ecuador_LandingCatch

